# Dissecting the mechanisms of a 5-year decline in antibiotic prescribing

**DOI:** 10.1101/2020.01.02.20016329

**Authors:** Stephen M. Kissler, R. Monina Klevens, Michael L. Barnett, Yonatan H. Grad

**Author notes:** Corresponding author: Yonatan H. Grad, MD, PhD, Department of Immunology and Infectious Diseases, Harvard T.H. Chan School of Public Health, 677 Huntington Avenue, Boston, MA 02115.

## Abstract

**Importance:** The mechanisms driving the recent decline in outpatient antibiotic prescribing are unknown.

**Objective:** To estimate the extent to which reductions in the number of antibiotic prescriptions filled per outpatient visit (stewardship) and reductions in the monthly rate of outpatient visits (observed disease) for infectious disease conditions each contributed to the decline in annual outpatient antibiotic prescribing rate in Massachusetts between 2011 and 2015.

**Design:** Outpatient medical and pharmacy claims from the Massachusetts All-Payer Claims Database were used to estimate rates of antibiotic prescribing and outpatient visits for 20 medical conditions and their contributions to the overall decline in antibiotic prescribing. Trends were compared to those in the National Ambulatory Medical Care Survey (NAMCS).

**Setting:** Outpatient visits in Massachusetts between January 2011 and September 2015.

**Participants:** 5,075,908 individuals with commercial health insurance or Medicaid in Massachusetts under the age of 65 and 495,515 patients included in NAMCS.

**Main outcomes and measures:** The number of antibiotic prescriptions avoided through reductions in observed disease and reductions in per-visit prescribing rate per medical condition.

**Results:** Between 2011 and 2015, the January antibiotic prescribing rate per 1,000 individuals in Massachusetts declined by 18.9% and the July antibiotic prescribing rate declined by 13.6%. The mean prescribing rate for children under 5 declined by 42.8% (95% CI 21.7%, 59.4%), principally reflecting reduced wintertime prescribing. The monthly rate of outpatient visits per 1,000 individuals in Massachusetts declined (*p* < 0.05) for respiratory infections and urinary tract infections. Nationally, visits for medical conditions that merit an antibiotic prescription also declined between 2010 and 2015. Of the estimated 358 antibiotic prescriptions per 1,000 individuals avoided over the study period in Massachusetts, 59% (95% CI 54%, 63%) were attributable to reductions in observed disease and 41% (95% CI 37%, 46%) to reductions in prescribing per outpatient visit.

**Conclusions and relevance:** The decline in antibiotic prescribing in Massachusetts was driven by a decline in observed disease and improved antibiotic stewardship, with a contemporaneous reduction in visits for conditions prompting antibiotics observed nationally. A focus on infectious disease prevention should be considered alongside antibiotic stewardship as a means to reduce antibiotic prescribing.

**Key points:** *Question:* How did the separate mechanisms of improved stewardship and reductions in observed disease contribute to a 5-year decline in outpatient antibiotic prescribing in Massachusetts from 2011-2015?

*Findings:* In an observational analysis of insurance claims, reduced monthly rates of outpatient visits for infectious conditions and reduced probability of prescribing an antibiotic per outpatient visit both contributed to the decline in antibiotic prescribing. An estimated 358 antibiotic prescriptions per 1,000 individuals were avoided over the study period through the two mechanisms, 211 of which were attributable to reductions in outpatient visits and 147 to reduced antibiotic prescribing per visit.

*Meaning:* Preventing the need for outpatient visits should be considered alongside antibiotic stewardship as a means of reducing antibiotic prescribing.

## Introduction

Reducing antibiotic use, particularly given the extent of inappropriate prescribing^1,2^, is a central public health goal in the effort to address the growing challenge of antibiotic resistance^3–5^. Between 2011 and 2015, the annual rate of filled outpatient antibiotic prescriptions to Massachusetts insurance holders under the age of 65 dropped by 17%, with similar declines reported across the United States^6,7^. However, the mechanisms driving this decline have been unclear. One mechanism could be improved antibiotic stewardship^8^ resulting in less inappropriate use. An alternative mechanism could be a reduction in antibiotic prescribing driven by a reduction in the incidence of “observed disease” —that is, the combined population-level rate of infectious illness and the probability of seeking heath care in response to illness. This mechanism would lead to a reduction in antibiotic use because fewer outpatient visits occur where prescribing—appropriate or inappropriate—is possible.

Reductions in observed disease could be attributed to increasing levels of self-care for minor illnesses, increasing use of telemedicine^9^, shifts in diagnostic coding, changing patient attitudes toward antibiotics, or to an authentic decline in disease, potentially driven by improved disease prevention, management, and vaccination^10,11^. A potentially important mechanism for disease prevention was the 2010 licensure of the 13-valent pneumococcal conjugate vaccine (PCV13),^12^ which produced substantial declines in pneumococcal disease^10,13^. If a reduction in observed disease is a significant contributor to declining antibiotic use, it would challenge the assumption that improving antibiotic stewardship is the key lever to decrease antibiotic use and control the rise of antibiotic resistance^14^.

The current literature examining declines in outpatient antibiotic prescribing in recent decades is mixed and has largely focused on changes in the degree of inappropriate prescribing^15–22^. A study of observed disease with respect to antibiotic prescribing for respiratory illnesses in the UK between 1994 and 2000 found that declines in observed respiratory disease accounted for a greater portion of the overall decline in prescribed antibiotics than improved stewardship^23^. In contrast, a study in the United States found that declines in antibiotic prescribing between 1995 and 2002 were primarily associated with improved stewardship, not with declines in observed disease^18^.

To address the gap in the understanding which mechanisms explain the declines in antibiotic prescribing, we used administrative claims from Massachusetts and a nationally representative dataset of outpatient office visits to quantify the respective contributions of improved stewardship and reduced observed disease to declining outpatient antibiotic prescribing. Massachusetts provides an ideal test case given that over 97% of residents have health insurance^24^ and the vast majority of medical insurance claims are reported to a centrally managed database^25^. This provides an unparalleled opportunity to define the population’s patterns of healthcare use.

## Methods

### Study sample and data sources

Medical and pharmacy claims data were obtained from the Massachusetts All-Payer Claims Database (APCD), 5^th^ edition^25^. The APCD contains insurance claims filed between 2011 and 2015 for 94% of Massachusetts residents under the age of 65 (Table S1). We examined trends through September 2015 because after October 2015, the ICD-10 diagnosis classification system was adopted nationally, unpredictably shifting observed prevalence of many conditions due to differences in coding^26^. We extracted all pharmacy claims for antibiotics using previously described methods^6^. We excluded pharmacy claims associated with individuals over the age of 65, since most of these individuals use traditional fee-for-service Medicare for their primary health insurance, and fee-for-service Medicare claims are not included in the APCD. Each antibiotic claim was associated with a unique patient identifier, the patient’s age, and the fill date^6^.

Outpatient physician encounters (‘visits’) were extracted from the APCD by pulling all medical claims associated with outpatient-associated Current Procedural Terminology (CPT) codes (Table S2). Each visit was associated with a unique patient identifier, the patient’s age, the service date, and up to seven ICD-9 coded diagnoses.

To examine national trends in observed disease, we extracted visits from the federal National Ambulatory Medicare Care Survey (NAMCS) dataset^27^ from 2002-2015. NAMCS is an annual, nationally representative survey of outpatient medical care in the United States. The dataset captures information on individual physician visits including up to three associated diagnoses. Each visit is weighted to allow extrapolation to national estimates.

### Defining medical conditions and associated prescriptions

Each diagnosis associated with outpatient visits in our study sample was mapped to one of the 20 conditions previously defined as associated with antibiotic prescribing^2^. For each visit, the diagnosis that ranked highest on the scale of antibiotic appropriateness^2^ was assigned as that visit’s primary diagnosis. We linked antibiotic prescriptions with visits on the basis of patient identifier and time between visits and prescription events ^16,28^. Each prescription could be linked to at most one visit; if a patient underwent multiple visits within the week preceding a filled antibiotic prescription, only the visit nearest to the prescription was linked. A prescription could fail to link with a visit if the associated visit did not qualify as an outpatient physician visit (e.g. the prescription was written during a dentistry visit or as a follow-up from a hospital stay), if the patient received the antibiotic without a formal outpatient visit (i.e., ‘phantom prescribing’^29^), if the prescription was filled more than one week after the visit, or if the patient used separate forms of insurance for the visit and the prescription fill, thereby associating the two claims with different patient IDs.

### Study outcomes and covariates

Our three primary outcomes were (a) the difference between the projected number of antibiotic prescriptions that would have been given over the study period in the absence of long-term trends in observed disease rates and (b) antibiotic prescribing rates as well as (c) the actual number of antibiotic prescriptions given over the study period. From these outcomes, we estimated the number of antibiotic prescriptions that were avoided due to changes in observed disease rates and/or changes in antibiotic prescribing rates. We measured these outcomes for each of the 20 potentially antibiotic meriting conditions independently. In a sensitivity analysis, we considered beneficiary age, sex, and county (Table S1) as covariates of observed disease and antibiotic prescribing rates.

We measured the monthly rate of antibiotic prescribing per 1,000 individual insurance enrollees, both overall and stratified by age, to assess longitudinal trends in prescribing. The denominator in each month consisted of the number of individuals enrolled in an insurance product for that entire month. In addition, we measured the annual physician visit rate per 1,000 people according to the NAMCS dataset for antibiotic-meriting and non-antibiotic-meriting conditions.

### Statistical analysis

For each of the 20 medical conditions, we calculated the monthly outpatient visit rate and the monthly number of associated antibiotic prescriptions per 1,000 individuals, where the denominator in both cases consisted of the number of individuals enrolled in an insurance product for the entire month. We then calculated the monthly rate of antibiotic prescribing per visit by dividing the monthly antibiotic prescriptions per 1,000 individuals by the monthly physician visit rate per 1,000 individuals. We used locally estimated scatterplot smoothing (LOESS) regression^30–32^ to visually highlight seasonal variation in antibiotic prescribing rates and outpatient visits.

Next, we projected the total number of antibiotic prescriptions that were avoided due to reductions in observed disease. For each of the 20 medical conditions, we calculated the slope of the monthly physician visit rate per 1,000 individuals using linear regression. We then projected the predicted monthly outpatient visit rate had there been no mean change in observed disease by adjusting the monthly incidence to have the same intercept but a regression slope over time of zero. Multiplying this adjusted monthly outpatient visit rate (visits per month per 1,000 individuals) by the actual monthly prescribing rate for that condition (prescriptions per visit) gave the projected number of monthly antibiotic prescriptions (prescriptions per month per 1,000 individuals) had there been no change in observed disease. We subtracted the actual number of monthly antibiotic prescriptions per 1,000 individuals for each condition from these projections to obtain the number of prescriptions per 1,000 individuals avoided due to reductions in observed disease. To assess the sensitivity of our findings to longitudinal changes in the demographic and geographic characteristics of the study population, we repeated the analysis using Poisson regression adjusted for month (seasonality), age, sex, and beneficiary county.

We applied the same procedure to the monthly per-visit prescribing rate to project the number of antibiotic prescriptions avoided due to improvements in stewardship. Multiplying the projected (zero-slope) monthly prescribing rate (prescriptions per visit) by the actual outpatient visit rate (visits per month per 1,000 individuals) and subtracting from the actual number of prescriptions filled over the study period yielded the number of prescriptions per 1,000 individuals that would have been given in the absence of long-term changes in stewardship.

To examine national trends in observed disease, we calculated the slope of the annual incidence of observed disease per 1,000 people for the conditions that always or sometimes merit an antibiotic (tiers 1 and 2) and the conditions that never merit an antibiotic (tier 3) from the NAMCS dataset using linear regression^2^. For the tier 1 and 2 conditions, we examined the possibility that the trend in annual observed disease incidence may have changed slope between 2002 and 2015 by fitting a piecewise-linear regression with a single breakpoint. We considered possible breakpoints in each six-month interval in 2002-2015 (January 1^st^ and July 1^st^ of each year) and chose the breakpoint that yielded the fit with the minimum sum of squared errors.

Antibiotic prescribing has a strong seasonal trend given the increased prevalence of viral respiratory infections in the winter^33^. To compare winter *vs*. summer declines in antibiotic prescribing, we fit a seasonal (harmonic) regression model^34^ with variable amplitude to the monthly rate of antibiotic prescriptions per 1,000 individuals:

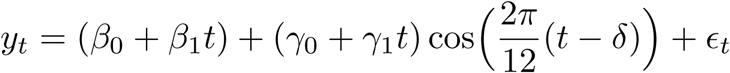

Here, *y*_*t*_ is the number of antibiotic prescriptions filled in month *t* per 1,000 individuals, *β*_*0*_ and *β*_*1*_ are the intercept and slope of the mean monthly prescribing rate per 1,000 individuals, *γ*_*0*_ is the amplitude of the seasonality at the start of the study period, *γ*_*1*_ is the monthly change in the seasonal amplitude, *δ* is a horizontal shift, and *ε*_*t*_ is an error term assumed to have mean zero and constant variance over time. A transformation of variables (see Supplemental Information) allowed us to compare the change over time of the peak (winter) *vs*. trough (summer) antibiotic prescribing rates. Code is available at https://github.com/skissler/disease-stewardship-ms.

## Results

The January antibiotic prescribing rate in Massachusetts declined from 78.5 prescriptions per 1,000 individuals in 2011 to 63.7 prescriptions per 1,000 individuals in 2015 (18.9% relative decrease), and the July antibiotic prescribing rate declined from 60.4 prescriptions per 1,000 individuals in 2011 to 52.2 prescriptions per 1,000 individuals in 2015 (13.6% relative decrease, Figure 1A). The mean monthly antibiotic prescribing rate declined by 42.8% (95% CI 21.7%, 59.4%) for children under the age of five (Figure 1B). Much of this decline is attributable to reduced wintertime prescribing: the winter monthly prescribing rate to children under the age of five declined at a rate of 1.2 prescriptions per 1,000 individuals per month (95% CI 0.89, 1.5) while the summer monthly prescribing rate declined at a rate of 0.30 prescriptions per 1,000 individuals per month (95% CI 0.012, 0.59) (Table S3, Figure S1).

**Figure 1.**
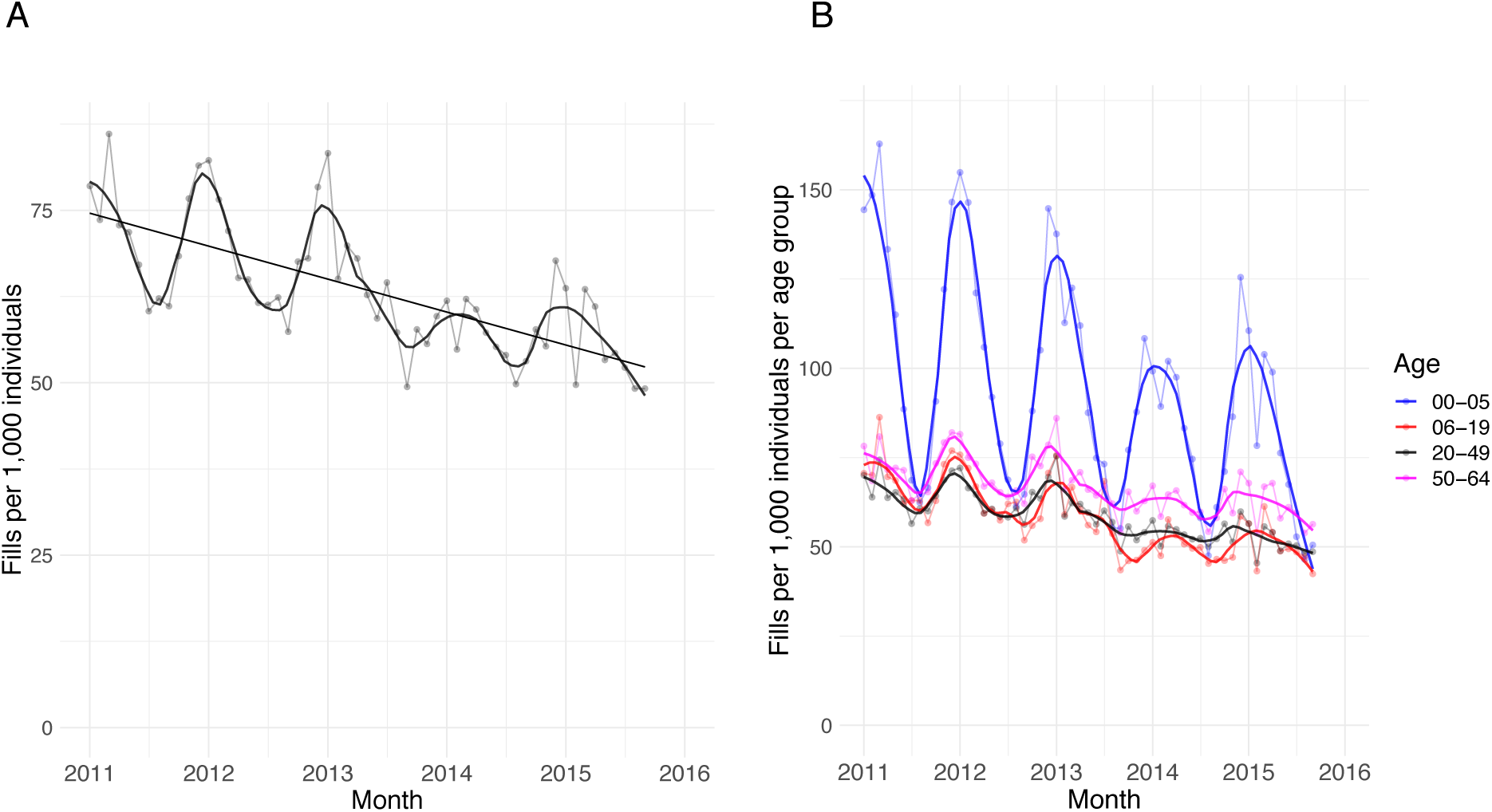
A: Monthly number of filled antibiotic prescriptions per 1,000 individuals in Massachusetts, 2011-2015 (points) with LOESS regression line (solid curve) using a span of 12 months. B: Monthly number of filled antibiotic prescriptions per 1,000 individuals in four age groups (points; 0-5 years, 6-19 years, 20-49 years, and 50-64 years) with LOESS regression line (solid curves) using a span of 12 months.

Declines in observed disease and improvements in stewardship together contributed to an estimated 358 (95% CI 324, 391) avoided antibiotic prescriptions per 1,000 individuals in Massachusetts between 2011 and 2015. An estimated 59% (95% CI 54%, 63%) of this decline was attributable to reductions in observed disease (Table 1). There were significant declines (regression slope *p*-value<0.05) in the monthly outpatient visit rates per 1,000 individuals for bronchitis/bronchiolitis, otitis media (suppurative and non-suppurative), pharyngitis, pneumonia (bacterial and viral), sinusitis, and urinary tract infections (Figures S2—S3). Figure 2 depicts the monthly visit rates for two conditions with significant declines (sinusitis and suppurative otitis media) and two conditions with non-significant declines (miscellaneous bacterial infections and skin, cutaneous and mucosal infections); the remaining conditions are shown in the Supplemental Information.

**Table 1.** Number of antibiotic prescriptions per 1,000 individuals filled for 20 medical conditions between January 2011 and September 2015, and the number of prescriptions per 1,000 individuals avoided through reductions in disease and improvements in prescribing. The 95% confidence intervals are given in parentheses. Negative values reflect that an increase in observed disease or a deterioration in stewardship contributed to an increase, rather than a reduction, in prescribing.

| Condition | Rx/1000 individuals | Rx/1000 avoided, disease | Rx/1000 avoided, prescribing |
| --- | --- | --- | --- |
| Sinusitis | 248 | 52.0 (34.0, 69.9) | 19.4 (15.5, 23.4) |
| Pharyngitis | 242 | 36.2 (23.8, 48.7) | 24.1 (19.1, 29.0) |
| Suppurative otitis media | 226 | 53.1 (35.4, 70.9) | 8.67 (5.76, 11.6) |
| Bronchitis/bronchiolitis | 52.4 | 19.4 (14.7, 24.2) | 11.9 (10.8, 13.0) |
| Viral upper respiratory infection | 84.8 | 11.6 (4.97, 18.3) | 12.7 (10.9, 14.4) |
| Pneumonia | 52.7 | 15.3 (10.2, 20.4) | 1.86 (0.984, 2.73) |
| Other skin cutaneous and mucosal conditions | 228 | 0.786 (-4.16, 5.73) | 15.0 (10.9, 19.0) |
| Asthma, allergy | 55.4 | 1.98 (0.0670, 3.90) | 12.8 (10.3, 15.4) |
| Skin cutaneous and mucosal infections | 179 | 2.70 (-5.05, 10.5) | 8.65 (3.57, 13.7) |
| Non-suppurative otitis media | 25.8 | 5.13 (4.09, 6.18) | 4.80 (3.95, 5.66) |
| Other genitourinary conditions | 119 | 3.86 (2.10, 5.61) | 5.27 (3.77, 6.77) |
| Urinary tract infections | 99.6 | 6.75 (4.60, 8.90) | 2.17 (0.761, 3.57) |
| Acne | 87.9 | -0.772 (-2.54, 0.994) | 9.33 (7.92, 10.7) |
| Gastrointestinal infections | 59.7 | 0.00310 (-1.16, 1.17) | 3.73 (2.90, 4.56) |
| Other respiratory conditions | 19.1 | 0.534 (0.168, 0.900) | 2.81 (2.09, 3.53) |
| Other gastrointestinal conditions | 48.1 | 0.308 (-0.454, 1.07) | 3.08 (2.43, 3.74) |
| Miscellaneous other infections | 10.4 | 0.0169 (-0.161, 0.194) | 1.14 (0.963, 1.32) |
| Miscellaneous bacterial infections | 65.4 | 2.40 (0.963, 3.83) | -1.33 (-2.77, 0.124) |
| Viral pneumonia | 0.851 | 0.461 (0.364, 0.558) | 0.0417 (0.0221, 0.0614) |
| Influenza | 1.98 | -0.886 (-1.90, 0.125) | 0.468 (0.188, 0.748) |
| **Total** | **1905** | **211 (180, 242)** | **147 (136, 157)** |

**Figure 2.**
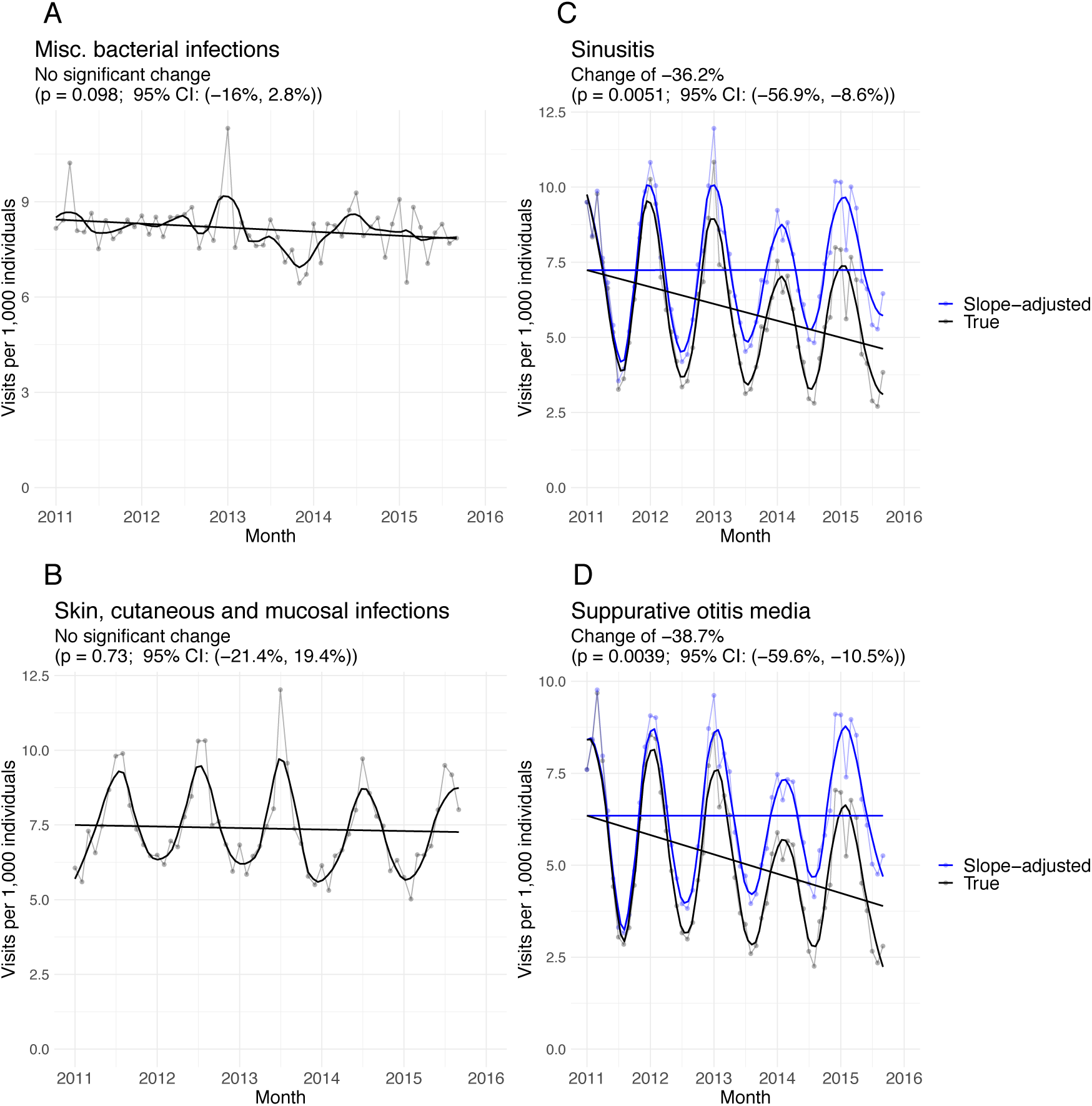
Monthly outpatient visit rate per 1,000 individuals in Massachusetts (points) for (A) miscellaneous bacterial infections, (B) skin, cutaneous and mucosal infections, (C) sinusitis, and (D) suppurative otitis media, with linear regression line (black, straight) and LOESS regression line using a span of 0.2 (black, curved). For conditions with a statistically significant decline in visit rate (p<0.05, subfigures C-D), the projected visit rate in the absence of a decline in observed disease is depicted in blue, with linear regression line (blue, straight) and LOESS regression line using a span of 0.2 (blue, curved).

The other 41% (95% CI 37%, 46%) of the overall decline in antibiotic prescribing was attributable to improved stewardship (Table 1). There were significant declines (regression slope *p-*value<0.05) in the monthly rate of antibiotic prescribing per outpatient visit for all conditions except influenza, miscellaneous bacterial infections, skin, cutaneous and mucosal infections, and urinary tract infections (Figures S4—S5). Figure 3 depicts the monthly per-visit antibiotic prescribing rates for two conditions with significant declines (sinusitis and suppurative otitis media) and two with non-significant declines (miscellaneous bacterial infections and skin, cutaneous and mucosal infections); the rest are depicted in the Supplemental Information. All findings were consistent with the demography- and geography-adjusted Poisson regression analysis (Table S4).

**Figure 3.**
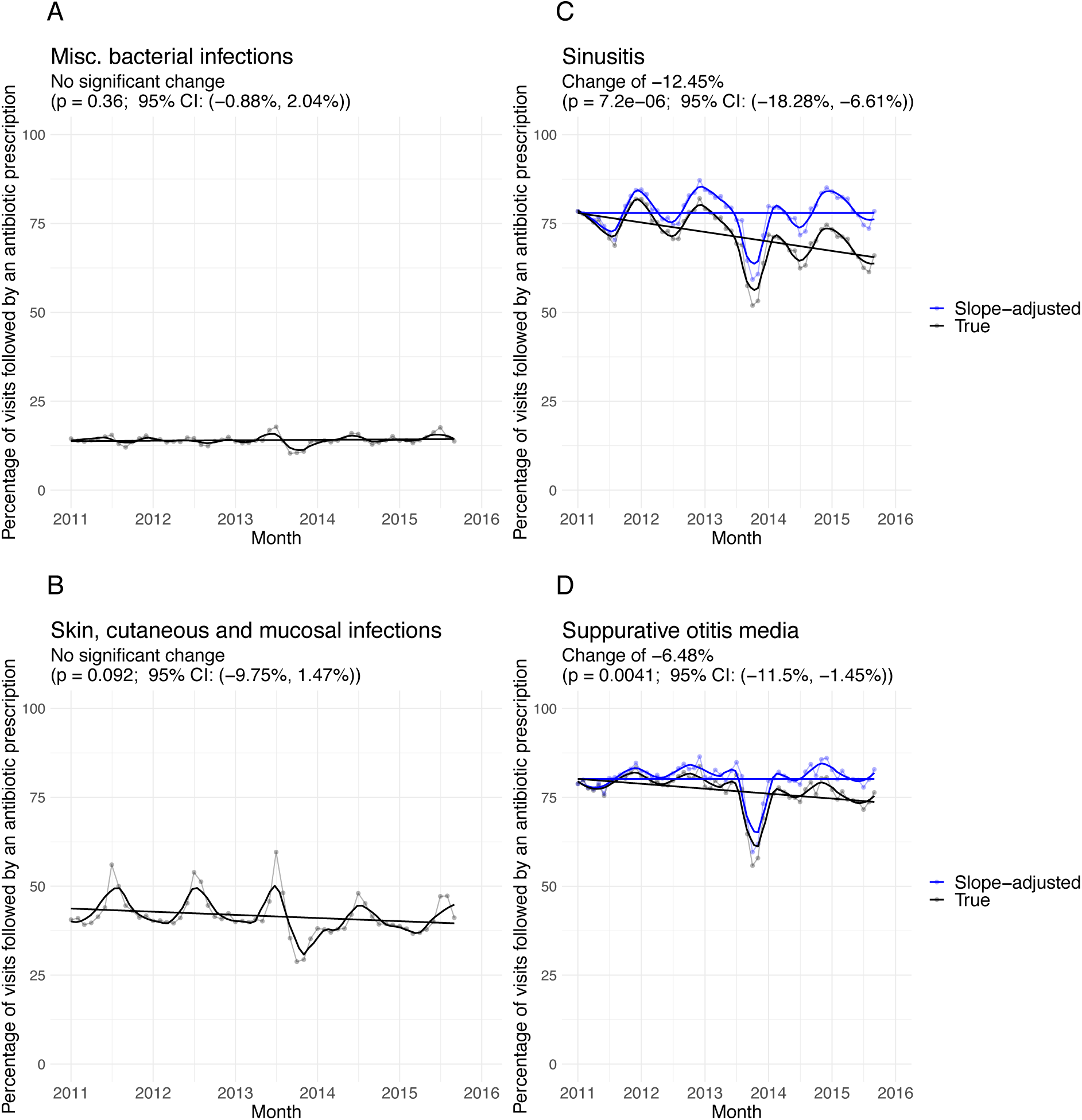
Monthly antibiotic prescribing rate per 1,000 individuals in Massachusetts for (A) miscellaneous bacterial infections, (B) skin, cutaneous, and mucosal infections, (C) sinusitis, and (D) suppurative otitis media, with linear regression line (black, straight) and LOESS regression line using a span of 0.2 (black, curved). For conditions with a statistically significant decline in prescribing rate (p<0.05, subfigures C-D), the projected prescribing rate in the absence of an improvement in stewardship is depicted in blue, with linear regression line (blue, straight) and LOESS regression line using a span of 0.2 (blue, curved).

We evaluated whether an increase in prescribing antibiotics without an associated visit (phantom prescribing^29^) could account for the decline in outpatient visits for infectious disease indications. However, the proportion of prescriptions in each month that could not be linked to an outpatient visit decreased by 5.2% from 48.4% (95% CI 46.4%, 50.4%) to 43.2% (95% CI 41.2%, 45.2%), inconsistent with the hypothesis that an increase in phantom prescribing could explain decreased visits for infectious disease indications (Figure S6).

To evaluate whether the declines in visits for these indications seen in Massachusetts reflect national trends, and thus whether the trend in decreased observed disease might explain the national decrease in antibiotic prescribing, we assessed the annual visit rates per 1,000 people for antibiotic-meriting indications (tiers 1 and 2) and non-antibiotic-meriting conditions (tier 3)^2^ between 2002 and 2015 using the NAMCS dataset^27^ (Figure 4). There was no significant change in the annual incidence per 1,000 people for the tier 3 conditions (slope *p*-value = 0.09), but the annual incidence of the tier 1 and 2 conditions declined by 22.2% (95% CI 10.1%, 32.8%). A piecewise-linear fit to the annual visit rate per 1,000 people for the tier 1 and 2 conditions reveals an especially pronounced decline in incidence between 2010 and 2015 (Figure 4).

**Figure 4.**
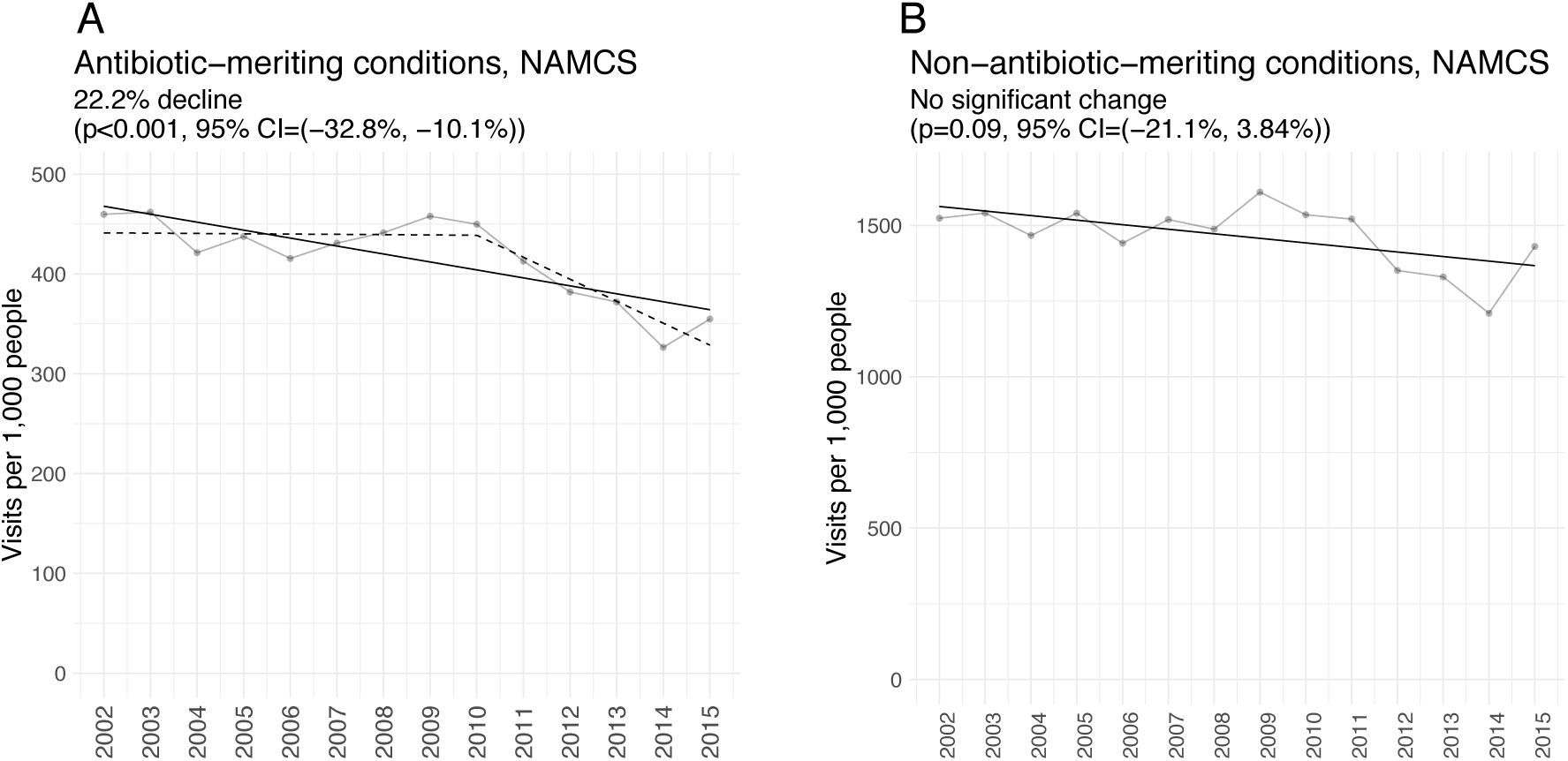
A: Annual rate of outpatient visits per 1,000 people in the United States associated with nine potentially antibiotic-meriting conditions (Tiers 1 and 2 in the classification by Fleming-Dutra *et al*. (2016)2) according to the NAMCS dataset, with linear regression line (solid) and piecewise-linear regression with maximum likelihood breakpoint in 2010 (dashed). B: Annual rate of outpatient visits per 1,000 people in the United States for the remaining 11 conditions that do not merit an antibiotic (Tier 3)2 according to the NAMCS dataset, with linear regression line. The linear trend remains statistically insignificant (slope *p*-value > 0.05) when the point at 2015 is excluded.

## Discussion

Improved stewardship and reduced rates of observed disease together contributed to reducing antibiotic prescriptions by an estimated 358 per 1,000 individuals in Massachusetts between January 2011 and September 2015. This reduction in prescribing was especially pronounced for wintertime pediatric prescribing. Stewardship improved significantly for 16 of the 20 of the conditions we considered, while rates of observed disease declined significantly for eight conditions, seven of which are associated with respiratory infections. The estimated number of prescriptions avoided through reduced disease were somewhat higher than the number avoided through improved stewardship. Our findings suggest that the prevention of diseases that prompt health care visits, especially respiratory diseases, should be considered alongside antibiotic stewardship as a way to reduce antibiotic prescribing.

The reasons behind the reduced rate of outpatient visits remain unclear. Much of the decline in observed disease might be attributable to respiratory illness prevented by the administration of PCV13 to children.^12,35^ The national decline in outpatient visits for antibiotic-meriting illnesses beginning in 2010 is consistent with this hypothesis, as are the declines in otitis media, sinusitis, bronchitis/bronchiolitis, pneumonia, and overall pediatric wintertime antibiotic prescribing in Massachusetts between 2011 and 2015. Evidence from randomized controlled trials indicate that pneumococcal vaccination reduces rates of antibiotic use^36^ and reduces the burden of antibiotic resistant infections.^37^ However, changing patient perceptions regarding antibiotics or mounting barriers to health care could also have led to an increasing tendency to self-care for minor illnesses, though, in this model, the unchanging rates of visits for other infectious indications would indicate that such trends in self-care are indication specific. Further studies are needed to distinguish between these possibilities.

Antibiotic stewardship played an integral role in reducing antibiotic prescribing in Massachusetts between 2011 and 2015. This is consistent with reports of successful antibiotic stewardship programs nationally^38^. We anticipate that these programs will remain key elements of strategies to reduce antibiotic prescribing and combat the rise of antibiotic resistance. However, there is a growing recognition that other strategies, including patient education and vaccination, will be required to achieve maximal reductions in antibiotic prescribing^39,40^. These preventative strategies have generally been regarded as secondary to stewardship. Our findings challenge the focus on stewardship by demonstrating that disease prevention can produce declines in antibiotic prescribing similar in magnitude to those achieved through stewardship, even when antibiotic prescribing was not an express target of those prevention measures.

Our findings are subject to a number of limitations. We only considered outpatient antibiotic prescriptions; however, these account for 80-90% of all antibiotic prescriptions^41^, so we believe we have captured the majority of prescriptions given in Massachusetts during the study period. Our analysis was limited to antibiotics that generated an insurance claim, omitting any that may have been obtained through less traditional routes. Also, we were unable to link a substantial minority of antibiotic claims with outpatient visits, and while these may represent a roughly random sample of all of the observed prescriptions, it is unclear if our conclusions about the linked prescriptions extend to the unlinked prescriptions. Finally, the geographic scope of our analysis was limited to one state, though we note that substantial declines in the incidence of antibiotic-meriting conditions occurred nationally during the same timeframe.

## Conclusions

Reductions in observed disease and improved stewardship similarly contributed to the decline in antibiotic prescribing in Massachusetts between 2011 and 2015. Our findings indicate that disease prevention could constitute a viable strategy for reducing antibiotic prescribing, on par with antibiotic stewardship. Further investigation into the causes of recent declines in respiratory disease could reveal new opportunities for intervention.

## Data Availability

Massachusetts APCD data are available from the Center for Health Information and Analysis[25]. NAMCS data are available from the Centers for Disease Control and Prevention[27]. Code for reproducing our analysis is available at https://github.com/skissler/disease-stewardship-ms.

## Supplemental materials and methods

**Supplementary Table S1.** Characteristics of the study population.

|  | 2011 | 2012 | 2013 | 2014 | 2015 |
| --- | --- | --- | --- | --- | --- |
| **Total (%)** | 4934998 (100) | 4948357 (100) | 5034557 (100) | 5075908 (100) | 5028609 (100) |
| **Age (%)** |  |  |  |  |  |
| 0-2 | 210180 (4.26) | 210635 (4.26) | 213167 (4.23) | 214768 (4.23) | 218055 (4.34) |
| 3-5 | 215702 (4.37) | 215846 (4.36) | 215429 (4.28) | 214502 (4.23) | 215260 (4.28) |
| 6-9 | 278700 (5.65) | 281085 (5.68) | 287062 (5.70) | 290453 (5.72) | 292336 (5.81) |
| 10-14 | 352291 (7.14) | 353145 (7.14) | 358745 (7.13) | 359267 (7.08) | 356914 (7.10) |
| 15-19 | 385998 (7.82) | 382153 (7.72) | 386114 (7.67) | 384040 (7.57) | 381791 (7.59) |
| 20-29 | 851266 (17.2) | 861933 (17.4) | 889151 (17.7) | 918331 (18.1) | 919999 (18.3) |
| 30-39 | 734711 (14.9) | 744018 (15.0) | 768042 (15.3) | 793797 (15.6) | 806076 (16.0) |
| 40-49 | 814690 (16.5) | 797461 (16.1) | 787840 (15.6) | 768113 (15.1) | 736272 (14.6) |
| 50-59 | 783253 (15.9) | 791136 (16.0) | 808720 (16.1) | 806320 (15.9) | 780585 (15.5) |
| 60-64 | 308207 (6.25) | 310945 (6.28) | 320287 (6.36) | 326317 (6.43) | 321321 (6.39) |
| **Female (%)** | 2533815 (51.3) | 2540025 (51.3) | 2580187 (51.2) | 2592417 (51.1) | 2565747 (51.0) |
| **County (%)** |  |  |  |  |  |
| Barnstable | 129418 (2.62) | 128307 (2.59) | 130480 (2.59) | 132150 (2.60) | 130801 (2.60) |
| Berkshire | 85526 (1.73) | 85036 (1.72) | 86488 (1.72) | 86756 (1.71) | 83391 (1.66) |
| Bristol | 367671 (7.45) | 369666 (7.47) | 376685 (7.48) | 384518 (7.58) | 382197 (7.60) |
| Dukes | 10118 (0.205) | 10033 (0.203) | 10474 (0.208) | 10055 (0.198) | 9507 (0.189) |
| Essex | 542536 (11.0) | 543679 (11.0) | 551771 (11.0) | 553425 (10.9) | 548099 (10.9) |
| Franklin | 48019 (0.973) | 46680 (0.943) | 46236 (0.918) | 45008 (0.887) | 41421 (0.824) |
| Hampden | 328800 (6.66) | 329314 (6.66) | 336465 (6.68) | 336704 (6.63) | 316489 (6.29) |
| Hampshire | 95061 (1.93) | 93086 (1.88) | 92553 (1.84) | 87918 (1.73) | 81196 (1.61) |
| Middlesex | 1111008 (22.5) | 1115564 (22.5) | 1139242 (22.6) | 1142107 (22.5) | 1129591 (22.5) |
| Nantucket | 7674 (0.156) | 7929 (0.160) | 8390 (0.167) | 8678 (0.171) | 9048 (0.180) |
| Norfolk | 489921 (9.93) | 492805 (9.96) | 503073 (9.99) | 502465 (9.90) | 491407 (9.77) |
| Plymouth | 354093 (7.18) | 352612 (7.13) | 357994 (7.11) | 358829 (7.07) | 352706 (7.01) |
| Suffolk | 512110 (10.4) | 522626 (10.6) | 542473 (10.8) | 564690 (11.1) | 574268 (11.4) |
| Worcester | 584610 (11.8) | 578549 (11.7) | 586219 (11.6) | 586057 (11.5) | 568673 (11.3) |
| Unknown | 268433 (5.44) | 272471 (5.51) | 266014 (5.28) | 276548 (5.45) | 309815 (6.16) |

**Supplementary Table S2.** CPT codes associated with outpatient visits^1^.

| **Code** | **Description** |
| --- | --- |
| 99201 - 99215 | Office or Other Outpatient Services |
| 99241 - 99245 | Office or Other Outpatient Consultation Services |
| 99354 - 99355 | Under Prolonged Service with Direct Patient Contact (Outpatient) |
| 99381 - 99397 | Preventive medicine |
| 99441 - 99444 | Telephone/online E/M |
| 99429 | Other preventive medicine services |
| 99499 | Other E/M Services |

### Measuring winter and summer declines in prescribing

To assess the difference between winter and summer declines in antibiotic prescribing in the under-five age group, we used nonlinear least squares regression as implemented in the `Non-linearModelFit[]` function in *Mathematica*^2^ to fit a sinusoid of the following form to the monthly number of antibiotic claims per 1,000 beneficiaries:

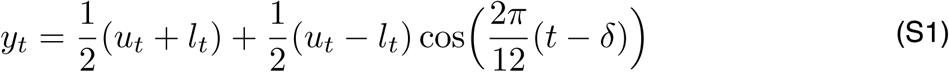

Here, *y*_*t*_ is the number of antibiotic claims per 1,000 beneficiaries in month *t, u*_*t*_ is the upper boundary of the sinusoid in month *t, l*_*t*_ is the lower boundary of the sinusoid in month *t*, and *δ* is the horizontal phase shift of the sinusoid in months. We assumed that the upper and lower boundaries of the sinusoid were linear trends defined by the linear equations

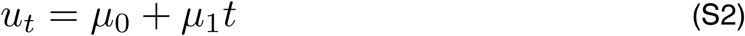

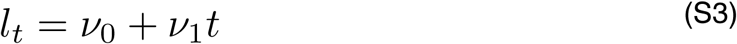

Here, *ε*_*0*_ and *ν*_*0*_ are the intercepts of the upper and lower boundaries of the sinusoid, respectively, and *ε*_*1*_ and *ν*_*1*_ are the slopes. This is equivalent to Eq. (1) in the main text under the following transformation of variables:

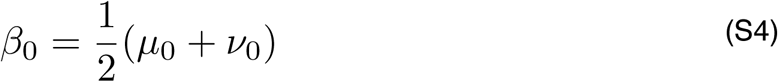

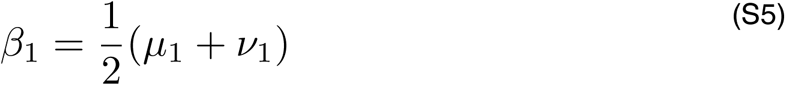

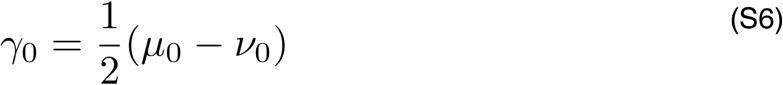

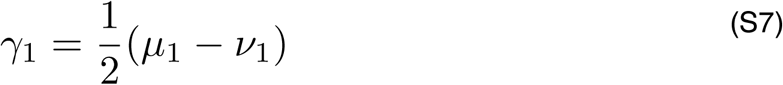

The fitting algorithm identified the maximum likelihood values for the parameters *μ*_*0*_, *μ*_*1*_, *ν*_*0*_, *ν*_*1*_, and *δ*. Table S1 gives the fitted values for those parameters with 95% confidence intervals. Figure S1 depicts the fitted sinusoid with the envelope defined by *u*_*t*_ and *l*_*t*_.

**Supplementary Table S3.**
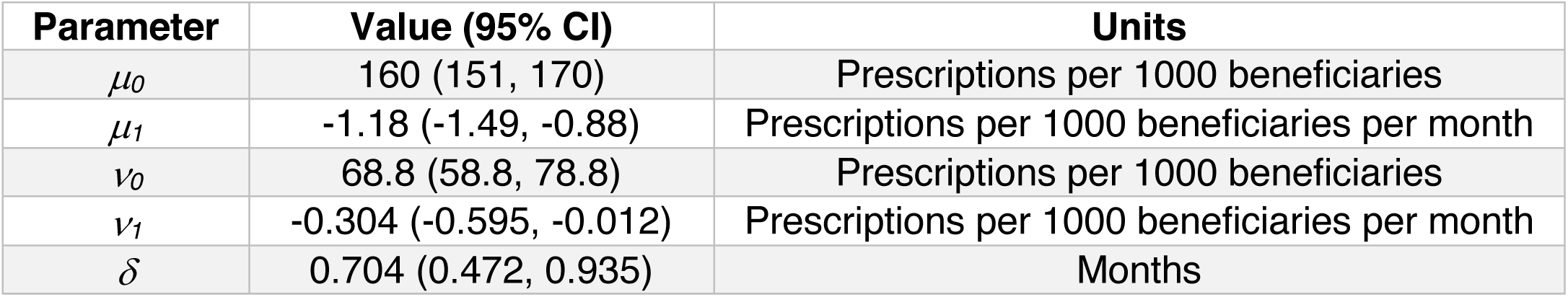
Fitted parameter values for the sinusoidal regression (Eq. S1-S3) to the monthly antibiotic prescribing rate per 1,000 beneficiaries under the age of 5.

**Supplementary Table S4.** Number of antibiotic prescriptions per 1,000 beneficiaries filled for 20 medical conditions between January 2011 and September 2015, and the number of prescriptions per 1,000 beneficiaries avoided through reductions in disease and improvements in prescribing using a Poisson regression model adjusted for month (seasonality), age, sex, and county. The 95% confidence intervals are given in parentheses. Negative values reflect that an increase in observed disease or a deterioration in stewardship contributed to an increase, rather than a reduction, in prescribing.

| **Condition** | **Rx/1000 beneficiaries** | **Rx/1000 avoided, disease** | **Rx/1000 avoided, prescribing** |
| --- | --- | --- | --- |
| **Sinusitis** | 248 | 35.9 (35.8, 36.1) | 16.9 (16.8, 17.0) |
| **Pharyngitis** | 242 | 26.9 (26.9, 27.0) | 22.5 (22.4, 22.6) |
| **Suppurative otitis media** | 226 | 39.0 (38.8, 39.1) | 7.77 (7.66, 7.88) |
| **Bronchitis/bronchiolitis** | 52.4 | 16.9 (16.8, 16.9) | 13.0 (12.9, 13.1) |
| **Viral upper respiratory infection** | 84.8 | 6.53 (6.50, 6.56) | 16.3 (16.2, 16.3) |
| **Skin cutaneous and mucosal infections** | 179 | 6.86 (6.79, 6.93) | 11.0 (10.8, 11.1) |
| **Pneumonia** | 52.7 | 13.3 (13.3, 13.4) | 0.952 (0.899, 1.00) |
| **Asthma, allergy** | 55.4 | 1.57 (1.55, 1.58) | 11.9 (11.8, 11.9) |
| **Other skin cutaneous and mucosal conditions** | 228 | 0.138 (0.0989, 0.177) | 13.4 (13.3, 13.5) |
| **Urinary tract infections** | 99.6 | 7.34 (7.27, 7.41) | 2.95 (2.87, 3.03) |
| **Non-suppurative otitis media** | 25.8 | 4.39 (4.37, 4.41) | 4.42 (4.38, 4.46) |
| **Other genitourinary conditions** | 119 | 2.53 (2.50, 2.56) | 5.90 (5.81, 5.98) |
| **Acne** | 87.9 | -0.853 (-0.896, -0.809) | 9.29 (9.21, 9.37) |
| **Other gastrointestinal conditions** | 48.1 | -0.470 (-0.483, -0.458) | 3.48 (3.42, 3.54) |
| **Gastrointestinal infections** | 59.7 | -1.04 (-1.06, -1.03) | 4.01 (3.94, 4.07) |
| **Other respiratory conditions** | 19.1 | 0.0786 (0.0694, 0.0877) | 2.52 (2.48, 2.56) |
| **Miscellaneous other infections** | 10.4 | 0.0203 (0.0140, 0.0266) | 1.19 (1.16, 1.22) |
| **Miscellaneous bacterial infections** | 65.4 | 1.73 (1.70, 1.75) | -1.10 (-1.16, -1.04) |
| **Viral pneumonia** | 0.851 | 0.505 (0.495, 0.516) | 0.0335 (0.0270, 0.0400) |
| **Influenza** | 1.98 | -0.788 (-0.791, -0.785) | 0.775 (0.745, 0.804) |
| **Total** | **1905** | **161 (159, 161)** | **147 (146, 148)** |

**Figure S1.**
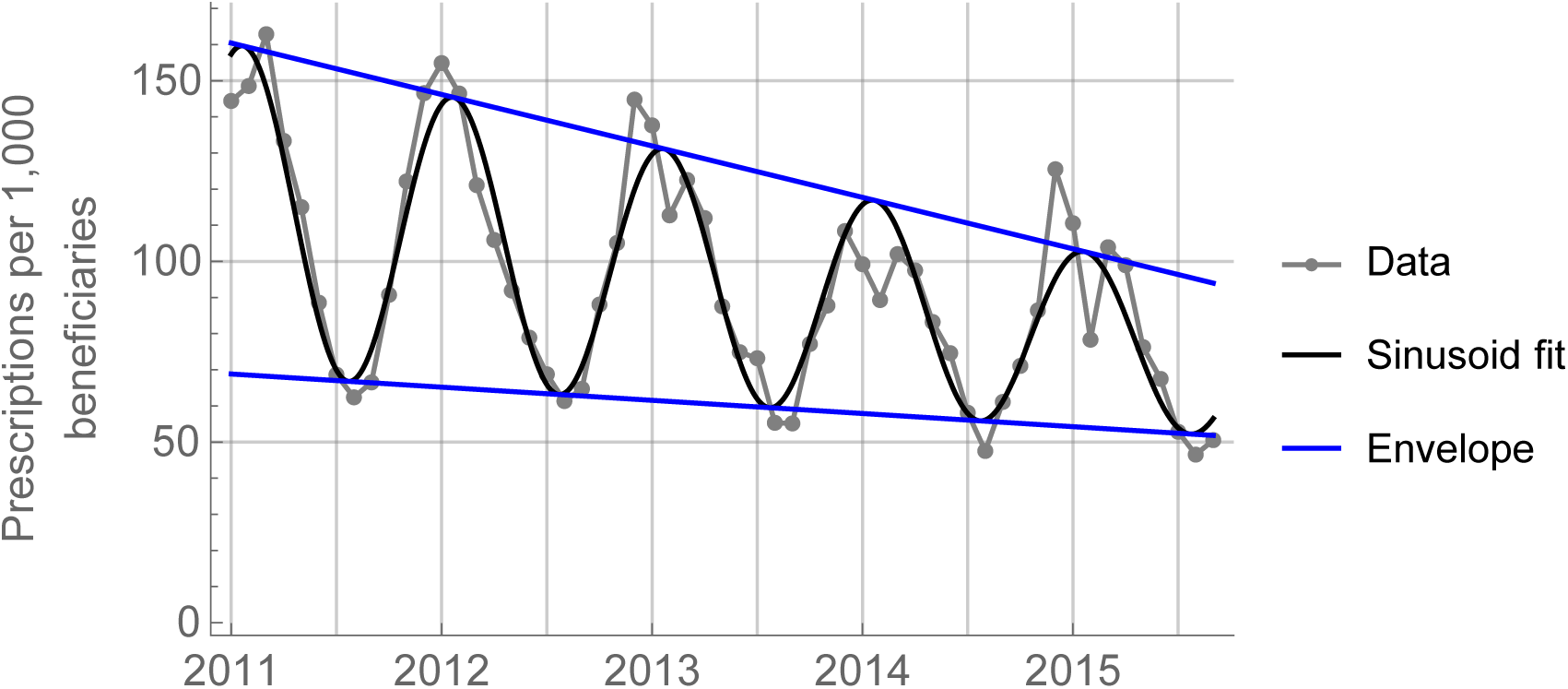
Monthly antibiotic prescribing rate per 1,000 beneficiaries under the age of 5 (grey, points) with maximum likelihood sinusoidal fit (black) and upper and lower envelope boundaries (*u*_*t*_ and *l*_*t*_, blue). The sinusoid is defined in Eq. S1 and the upper and lower boundaries are defined in Eq. S2-S3. Parameter values for the sinusoid are given in Table S3.

**Figure S2.**
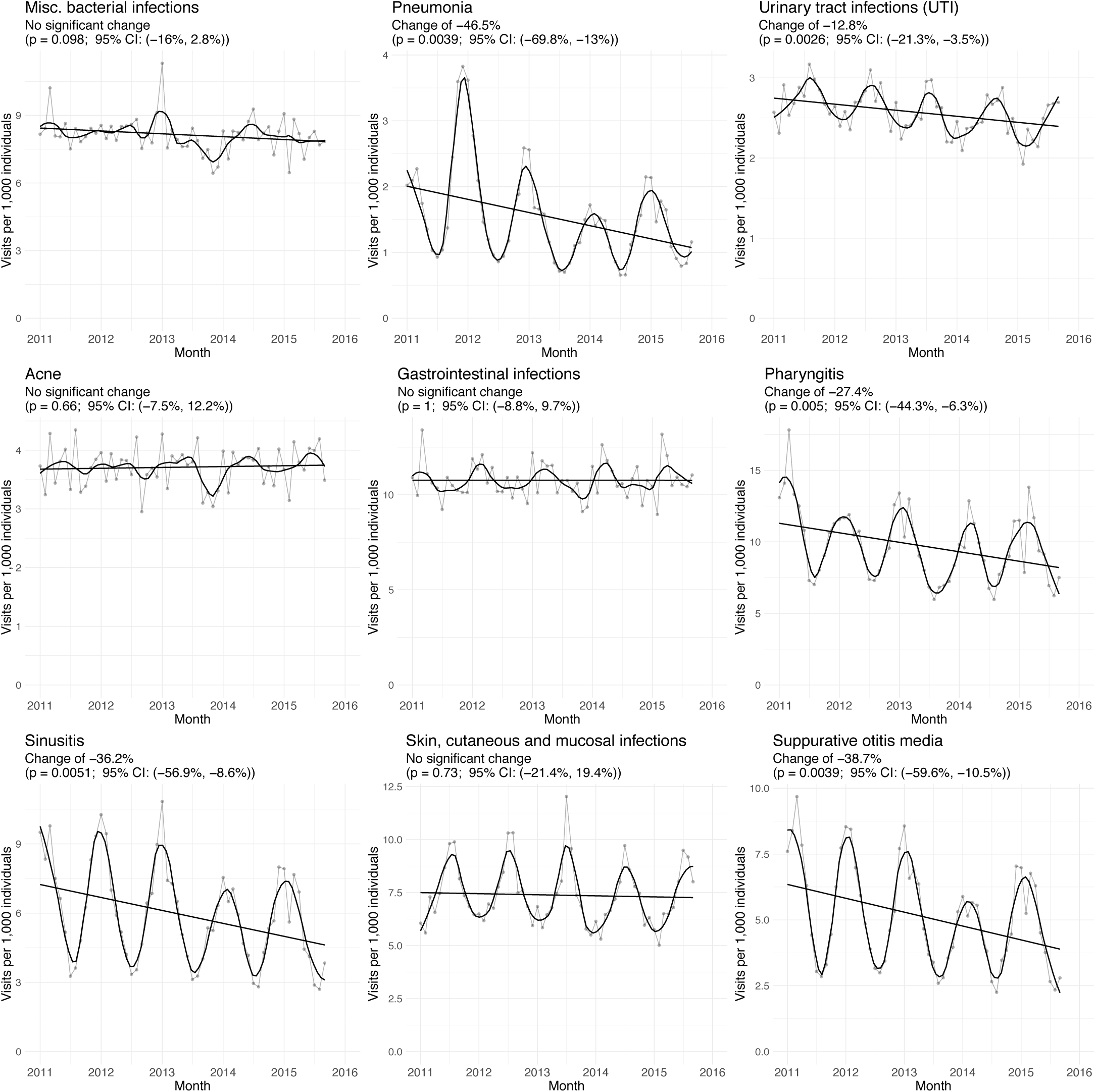
Monthly rate of observed disease per 1,000 beneficiaries for the Tier 1 and 2 conditions^3^.

**Figure S3.**
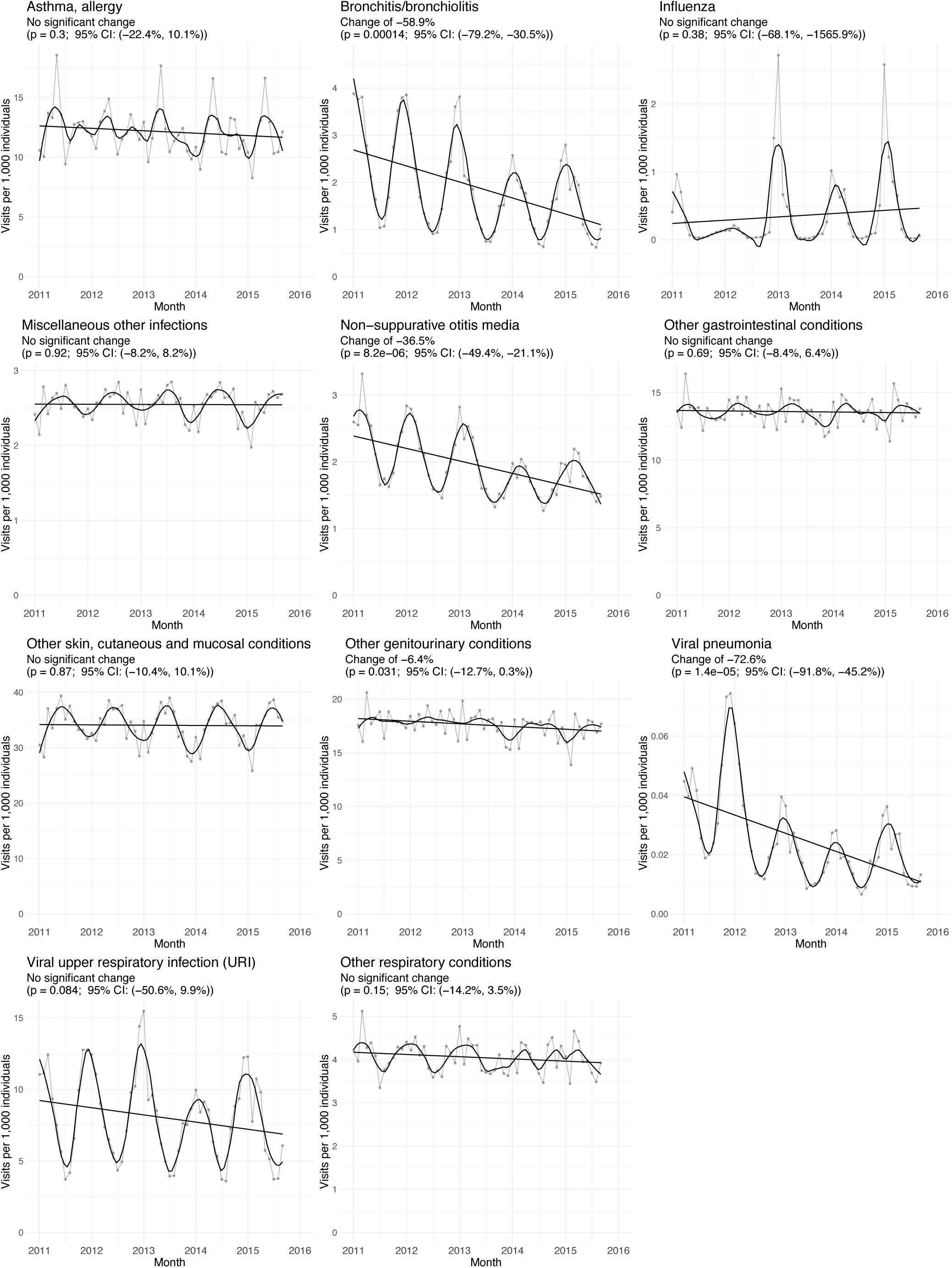
Monthly rate of observed disease per 1,000 beneficiaries for the Tier 3 conditions^3^.

**Figure S4.**
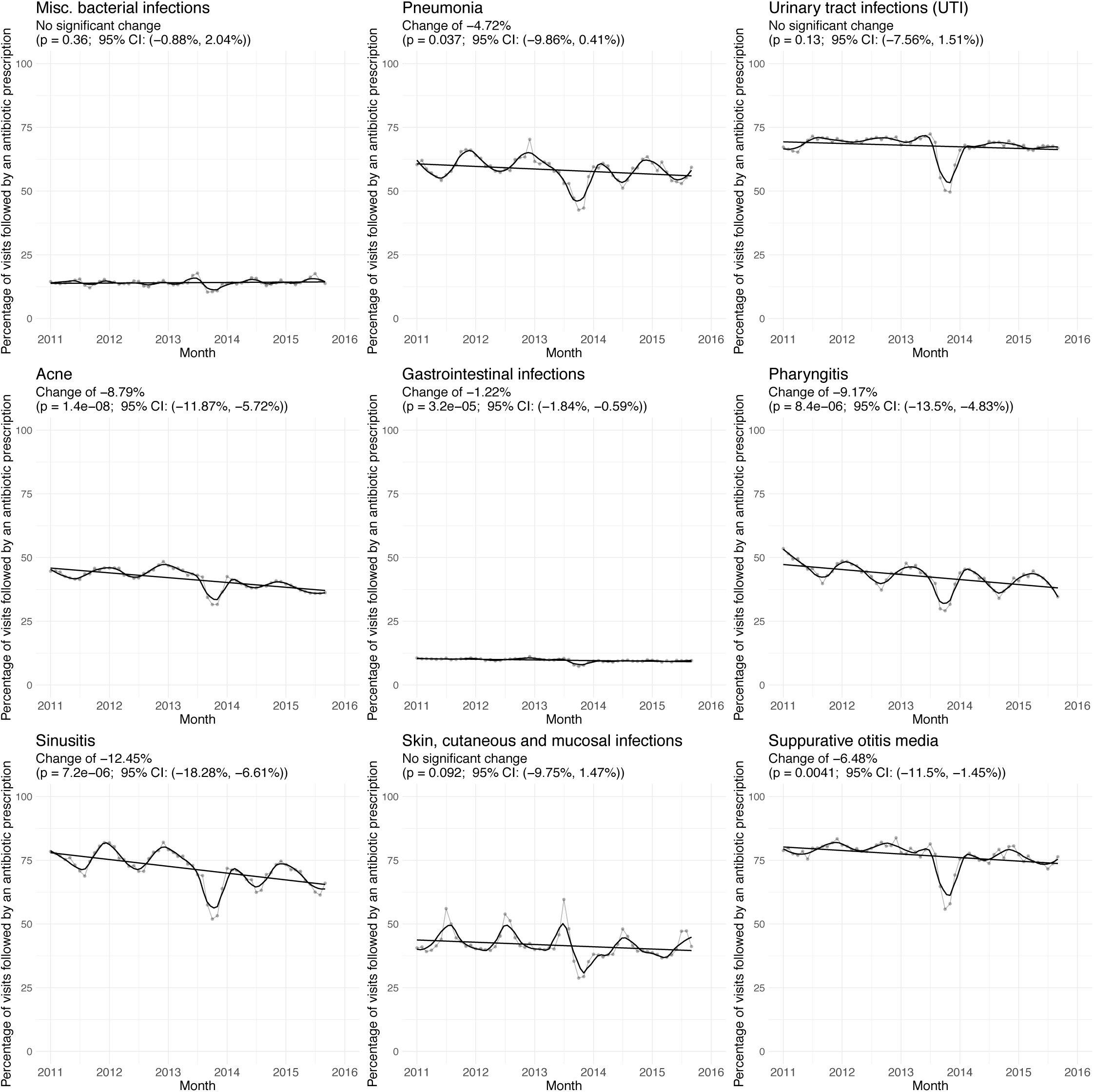
Monthly prescribing rate per physician visit for the Tier 1 and 2 conditions^3^.

**Figure S5.**
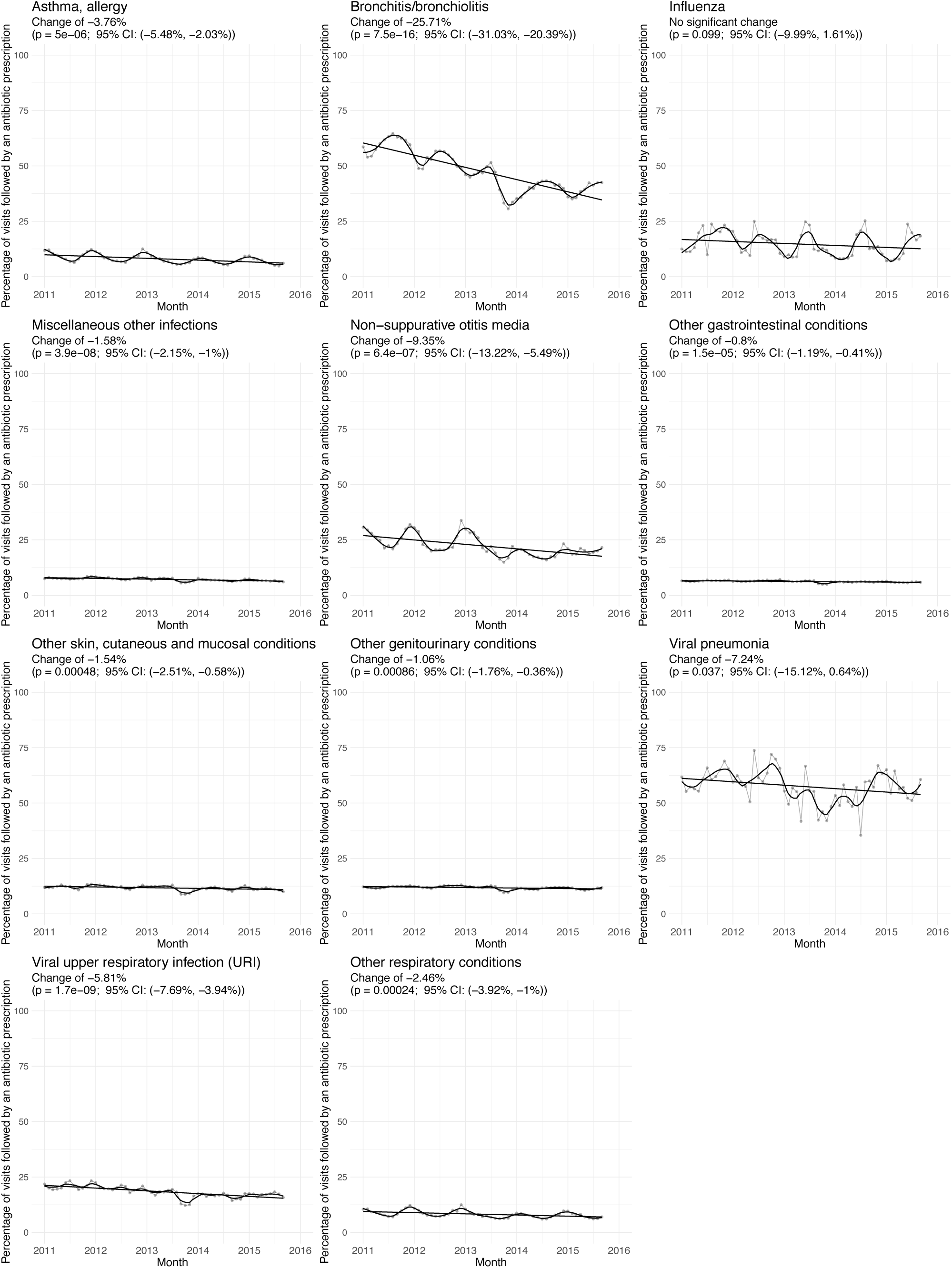
Monthly prescribing rate per physician visit for the Tier 3 conditions^3^.

**Figure S6.**
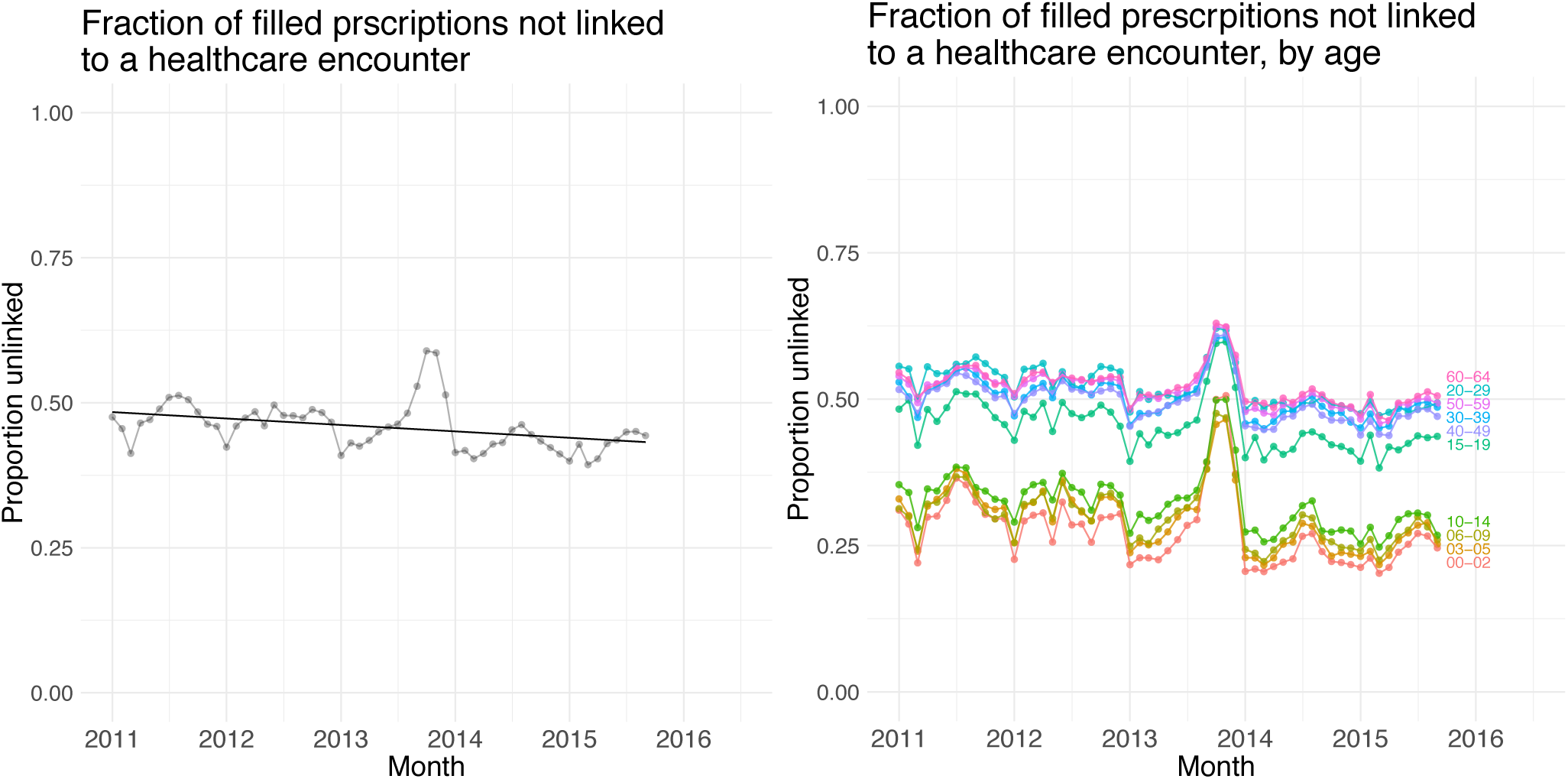
A: Monthly fraction of prescriptions not linked with an outpatient healthcare encounter. B: Monthly fraction of prescriptions not linked with an outpatient healthcare encounter, by age group.

**Figure S7.**
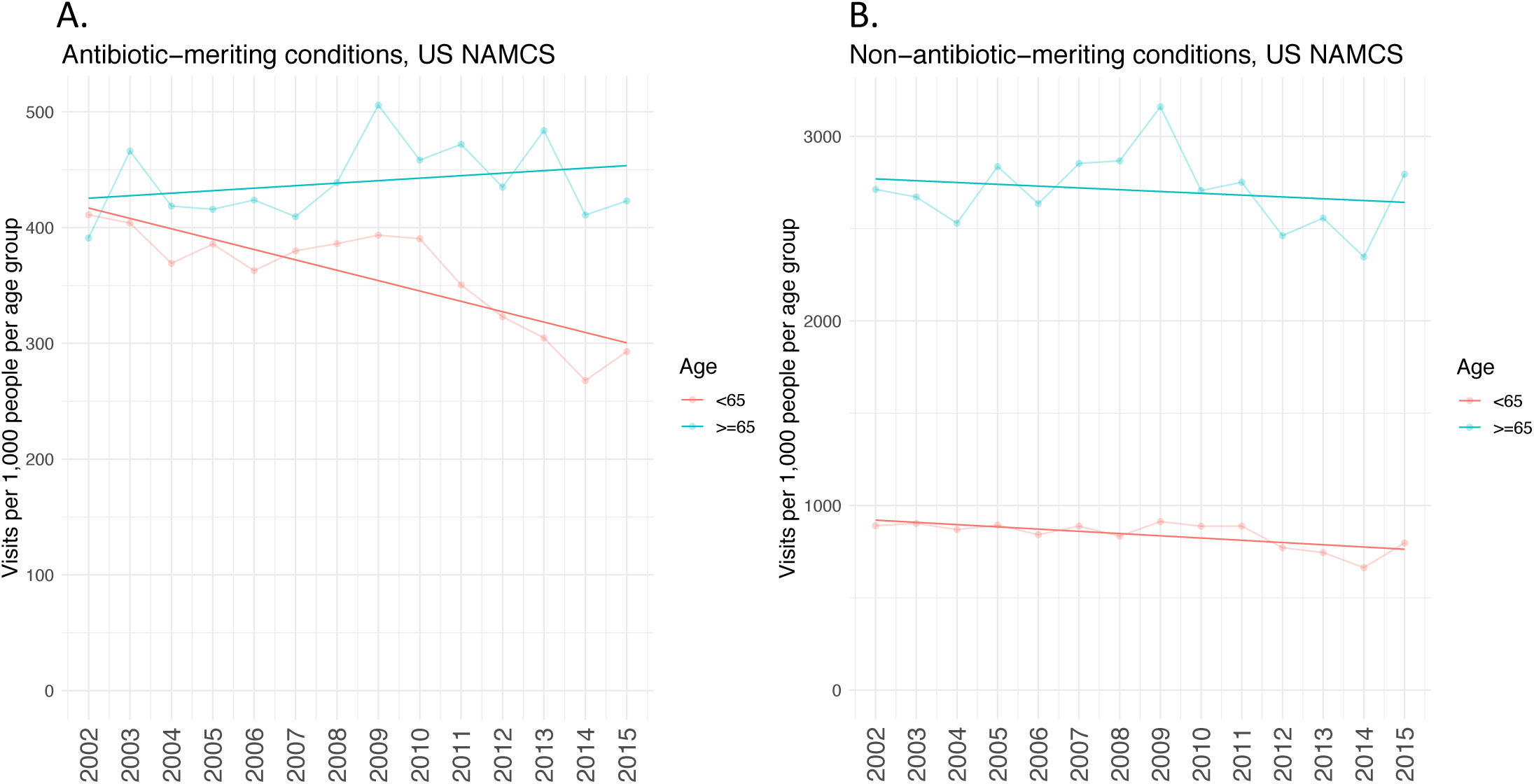
A. Annual physician visit rate for antibiotic-meriting conditions (tiers 1 and 2) according to the NAMCS dataset, separated by age group. The *p*-values for the <65 and >=65 trend lines are 0.000173 and 0.341, respectively. B. Annual physician visit rate for the non-antibiotic-meriting conditions (tier 3) according to the NAMCS dataset, separated by age group. The *p*-values for the <65 and >=65 trend lines are 0.0058 and 0.487, respectively.

---

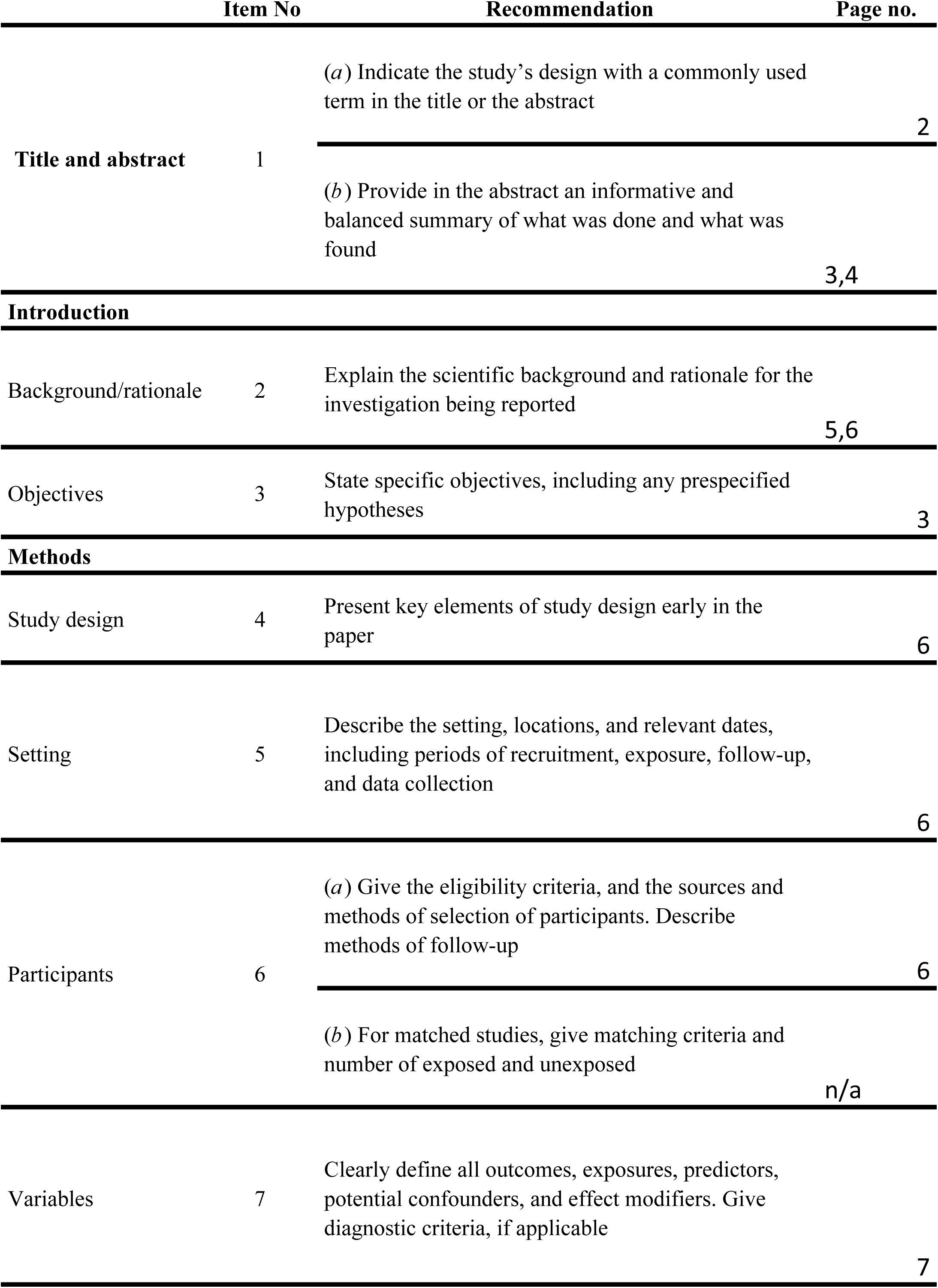
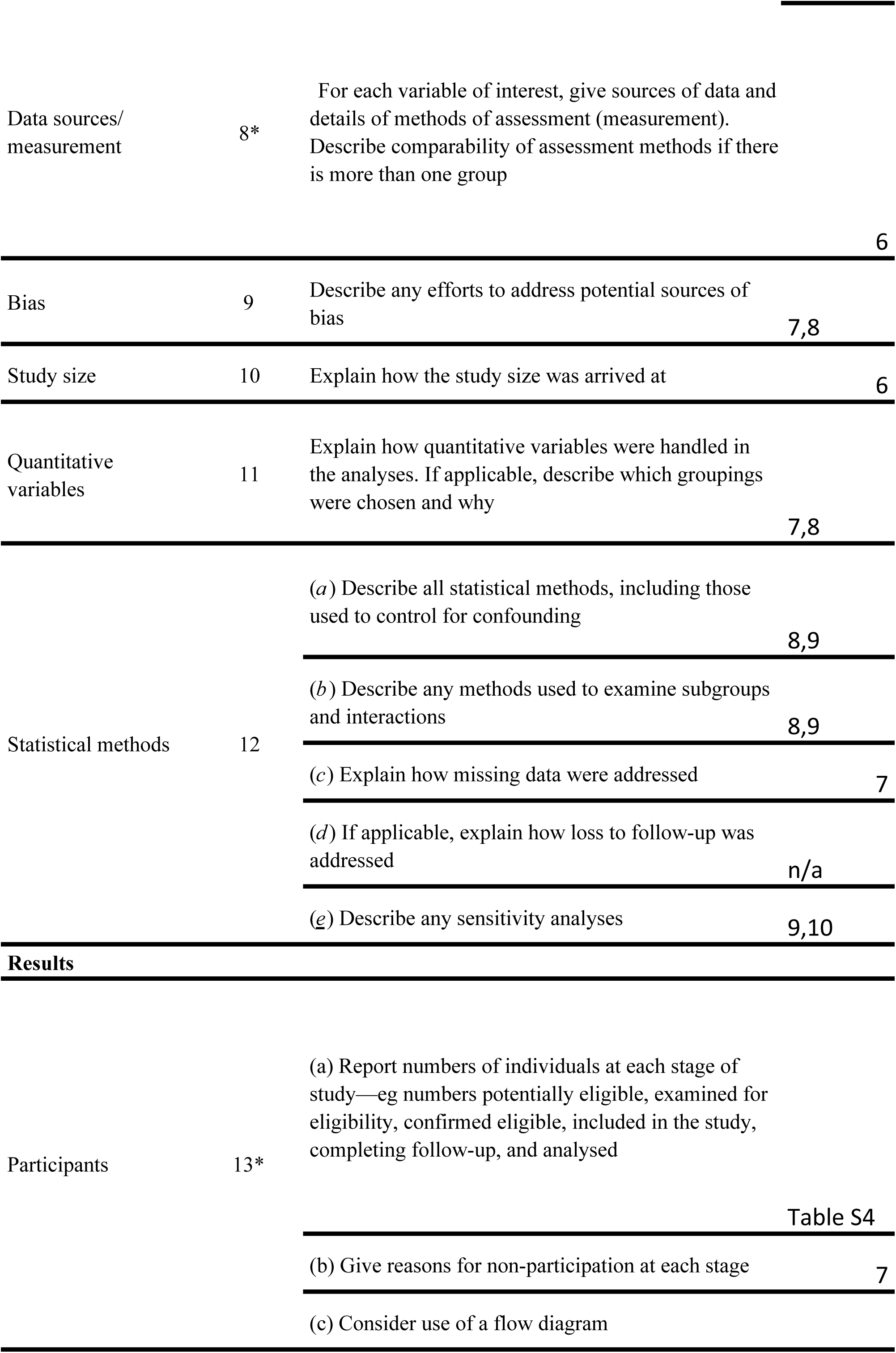
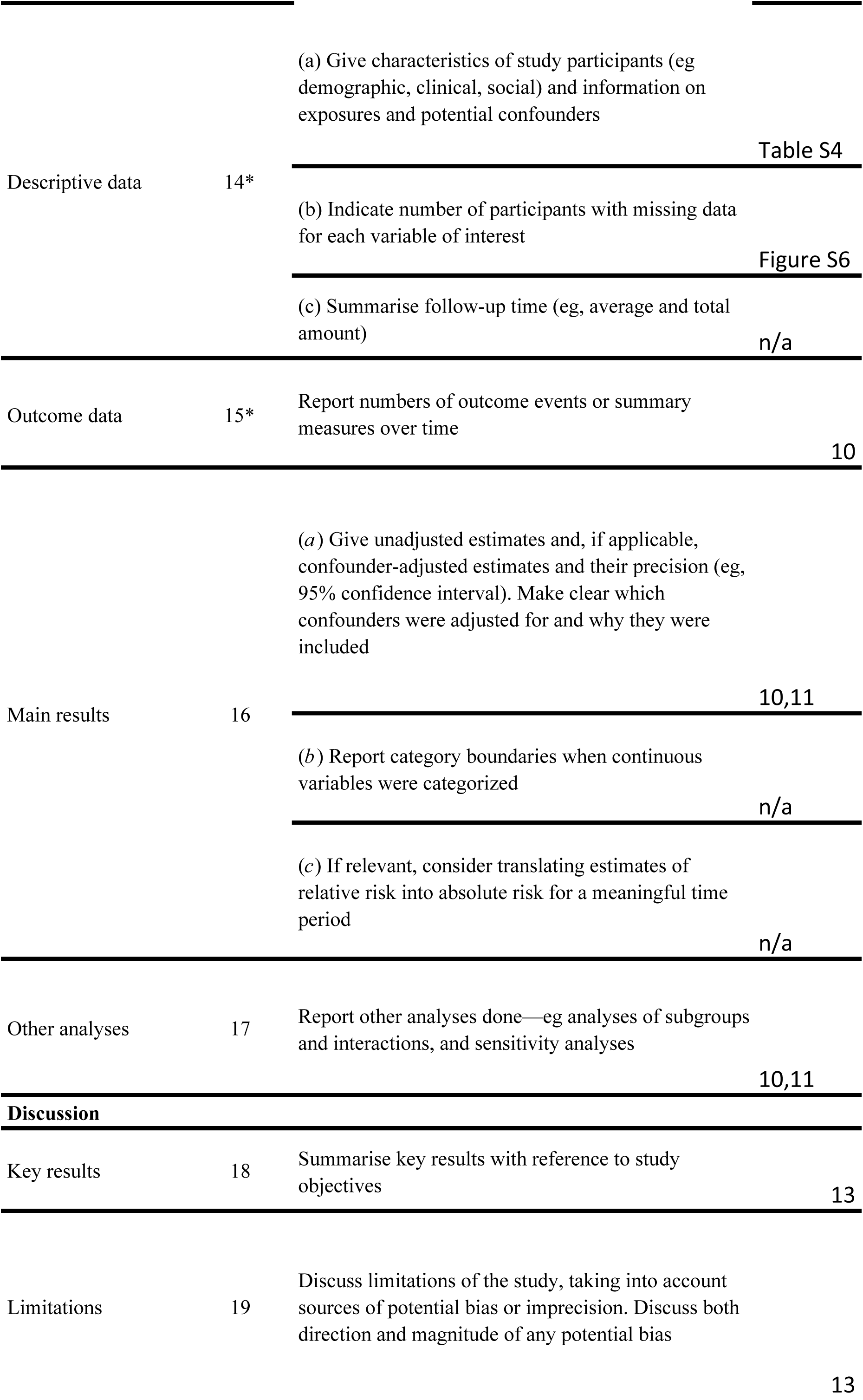
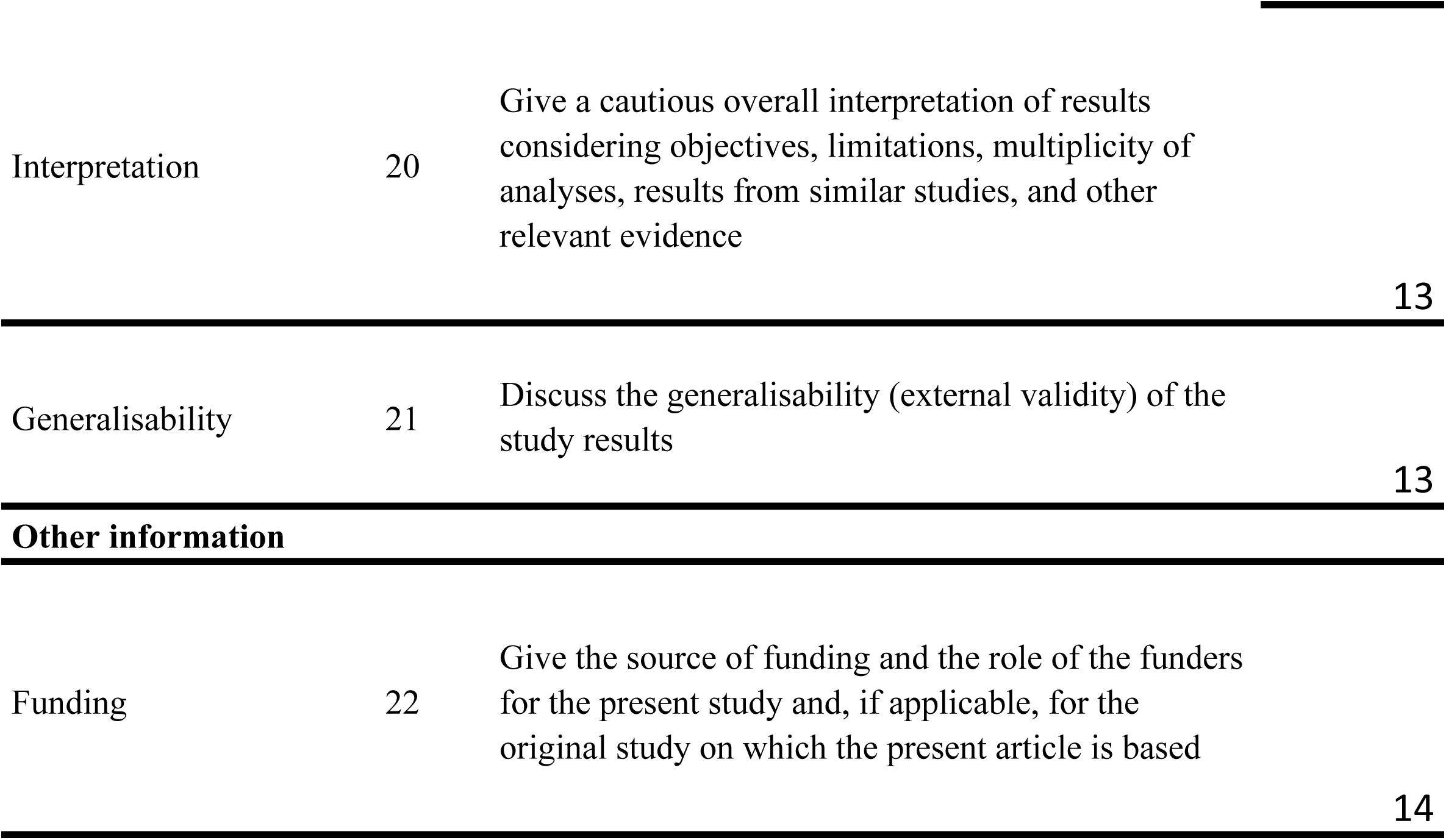

## References

1. Chua K-P, Fischer MA, Linder JA. Appropriateness of outpatient antibiotic prescribing among privately insured US patients: ICD-10-CM based cross sectional study. BMJ. January 2019:k5092. doi:10.1136/bmj.k5092

2. Fleming-Dutra KE, Hersh AL, Shapiro DJ, et al. Prevalence of inappropriate antibiotic prescriptions among us ambulatory care visits, 2010-2011. JAMA - J Am Med Assoc. 2016;315(17):1864–1873. doi:10.1001/jama.2016.4151

3. Sanchez G V., Fleming-Dutra KE, Roberts RM, Hicks LA. Core Elements of Outpatient Antibiotic Stewardship. MMWR Recomm Reports. 2016;65(6):1–12. doi:10.15585/mmwr.rr6506a1

4. Centers for Disease Control and Prevention. Antibiotic prescribing and use. https://www.cdc.gov/antibiotic-use/index.html. Published 2019. Accessed July 8, 2019.

5. World Health Organization. Antimicrobial resistance. https://www.who.int/antimicrobial-resistance/en/. Published 2019. Accessed July 8, 2019.

6. Klevens RM, Caten E, Olesen SW, DeMaria A, Troppy S, Grad YH. Outpatient Antibiotic Prescribing in Massachusetts, 2011–2015. Open Forum Infect Dis. 2019;6(5). doi:10.1093/ofid/ofz169

7. Blue Cross Blue Shield. Antibiotic prescription fill rates declining in the U.S. 2017.

8. Centers for Disease Control and Prevention. Antibiotic Prescribing and Use: Be Antibiotics Aware.

9. Barnett ML, Ray KN, Souza J, Mehrotra A. Trends in Telemedicine Use in a Large Commercially Insured Population, 2005-2017. JAMA - J Am Med Assoc. 2018;320(20):2147–2149. doi:10.1001/jama.2018.12354

10. Pilishvili T. Impact of PCV13 on Invasive Pneumococcal Disease (IPD) Burden and the Serotype Distribution in the U.S. Atlanta, GA; 2018.

11. Vadlamudi NK, Chen A, Marra F. Impact of the 13-Valent Pneumococcal Conjugate Vaccine Among Adults : A Systematic Review and Meta-analysis. 2019;69:34–49. doi:10.1093/cid/ciy872

12. Centers for Disease Control and Prevention. Licensure of a 13-Valent Pneumococcal Conjugate Vaccine (PCV13) and Recommendations for Use Among Children --- Advisory Committee on Immunization Practices (ACIP), 2010. Atlanta, GA; 2010.

13. Bruhn CAW, Hetterich S, Schuck-Paim C, et al. Estimating the population-level impact of vaccines using synthetic controls. Proc Natl Acad Sci U S A. 2017;114(7):1524–1529. doi:10.1073/pnas.1612833114

14. The White House. National Strategy for Combating Antibiotic-Resistant Bacteria. Washington, DC; 2014.

15. Centers for Disease Control and Prevention. Office-Related Antibiotic Prescribing for Persons Aged ≤14 Years --- United States, 1993--1994 to 2007--2008. Atlanta, GA; 2011.

16. Vaz LE, Kleinman KP, Raebel MA, et al. Recent Trends in Outpatient Antibiotic Use in Children. Pediatrics. 2014;133(3):375–385. doi:10.1542/peds.2013-2903

17. Suda KJ, Roberts RM, Hunkler RJ, Taylor TH. Antibiotic prescriptions in the community by type of provider in the United States, 2005-2010. J Am Pharm Assoc. 2016;56(6):621–626.e1. doi:10.1016/j.japh.2016.08.015

18. Roumie CL, Halasa NB, Grijalva CG, et al. Trends in antibiotic prescribing for adults in the United States - 1995 to 2002. J Gen Intern Med. 2005;20(8):697–702. doi:10.1111/j.1525-1497.2005.0148.x

19. Durkin MJ, Jafarzadeh SR, Hsueh K, et al. Outpatient antibiotic prescription trends in the United States: A national cohort study. Infect Control Hosp Epidemiol. 2018;39(5):584–589. doi:10.1017/ice.2018.26

20. Hicks LA, Bartoces MG, Roberts RM, et al. US outpatient antibiotic prescribing variation according to geography, patient population, and provider specialty in 2011. Clin Infect Dis. 2015;60(9):1308–1316. doi:10.1093/cid/civ076

21. Schmidt ML, Spencer MD, Davidson LE. Patient, provider, and practice characteristics associated with inappropriate antimicrobial prescribing in ambulatory practices. Infect Control Hosp Epidemiol. 2018;39(3):307–315. doi:10.1017/ice.2017.263

22. Barlam TF, Soria-Saucedo R, Cabral HJ, Kazis LE. Unnecessary antibiotics for acute respiratory tract infections: Association with care setting and patient demographics. Open Forum Infect Dis. 2016;3(1):1–7. doi:10.1093/ofid/ofw045

23. Ashworth M, Latinovic R, Charlton J, Cox K, Rowlands G, Gulliford M. Why has antibiotic prescribing for respiratory illness declined in primary care? A longitudinal study using the General Practice Research Database. J Public Health (Bangkok). 2004;26(3):268–274. doi:10.1093/pubmed/fdh160

24. Berchick ER, Barnett JC, Upton RD. Current Population Reports, P60-267 (RV), Health Insurance Coverage in the United States: 2018. Washington, DC; 2019. doi:10.1093/ije/dys118

25. Center for Health Information and Analysis. Massachusetts All-Payer Claims Database.

26. Khera R, Dorsey KB, Krumholz HM. Transition to the ICD-10 in the United States. JAMA. 2018;320(2):133. doi:10.1001/jama.2018.6823

27. National Center for Health Statistics. Ambulatory Health Care Data. https://www.cdc.gov/nchs/ahcd/index.htm. Published 2019. Accessed July 31, 2019.

28. Mundkur ML, Franklin J, Huybrechts KF, et al. Changes in Outpatient Use of Antibiotics by Adults in the United States, 2006–2015. Drug Saf. 2018;41(12):1333–1342. doi:10.1007/s40264-018-0697-4

29. Riedle BN, Polgreen LA, Cavanaugh JE, Schroeder MC, Polgreen PM. Phantom Prescribing: Examining the Frequency of Antimicrobial Prescriptions Without a Patient Visit. Infect Control Hosp Epidemiol. 2017;38(3):273–280. doi:10.1017/ice.2016.269

30. Cleveland WS, Devlin SJ. Locally weighted regression: An approach to regression analysis by local fitting. J Am Stat Assoc. 1988;83(403):596–610. doi:10.1080/01621459.1988.10478639

31. R Core Team. R: A language and environment for statistical computing. 2018.

32. Wickham H. Ggplot2: Elegant Graphics for Data Analysis. New York: Springer-Verlag; 2016.

33. Fisman D. Seasonality of viral infections: Mechanisms and unknowns. Clin Microbiol Infect. 2012;18(10):946–954. doi:10.1111/j.1469-0691.2012.03968.x

34. Brockwell PJ, Davis RA. Introduction to Time Series and Forecasting. (Brockwell PJ, Davis RA, eds.). New York, NY: Springer New York; 2002. doi:10.1007/b97391

35. Centers for Disease Control and Prevention. MMWR 2010. Vol 59. Atlanta, GA; 2010.

36. Buckley BS, Henschke N, Bergman H, et al. Impact of vaccination on antibiotic usage: a systematic review and meta-analysis. Clin Microbiol Infect. 2019. doi:10.1016/j.cmi.2019.06.030

37. Klugman KP, Black S. Impact of existing vaccines in reducing antibiotic resistance: Primary and secondary effects. Proc Natl Acad Sci. 2018;115(51):12896–12901. doi:10.1073/pnas.1721095115

38. Grammatico-Guillon L, Abdurrahim L, Shea K, Astagneau P, Pelton S. Scope of Antibiotic Stewardship Programs in Pediatrics. Clin Pediatr (Phila). 2019;58(11-12):1291–1301. doi:10.1177/0009922819852985

39. Hersh AL, Fleming-Dutra KE. Vaccines and Outpatient Antibiotic Stewardship. Pediatrics. 2017;140(3):e20171695. doi:10.1542/peds.2017-1695

40. Sevilla JP, Bloom DE, Cadarette D, Jit M, Lipsitch M. Toward economic evaluation of the value of vaccines and other health technologies in addressing AMR. Proc Natl Acad Sci U S A. 2018;115(51):12911–12919. doi:10.1073/pnas.1717161115

41. Suda KJ, Hicks LA, Roberts RM, Hunkler RJ, Matusiak LM, Schumock GT. Antibiotic Expenditures by Medication, Class, and Healthcare Setting in the United States, 2010-2015. Clin Infect Dis. 2018;66(2):185–190. doi:10.1093/cid/cix773

## Bibliography

1. American Academy of Professional Coders. AAPC Coder. CPT Codes. https://coder.aapc.com/cpt-codes. Published 2019. Accessed November 21, 2019.

2. Wolfram Research Inc. Mathematica. 2019.

3. Fleming-Dutra KE, Hersh AL, Shapiro DJ, et al. Prevalence of inappropriate antibiotic prescriptions among us ambulatory care visits, 2010-2011. JAMA - J Am Med Assoc. 2016;315(17):1864–1873. doi:10.1001/jama.2016.4151

